# Feasibility of and Experience with Telehealth Based Patient Self-referral for COVID-19 Monoclonal Antibody Therapy

**DOI:** 10.1101/2022.07.07.22277363

**Authors:** Ishaan Gupta, Sophia Purekal, Yahya Shaikh, Henry J. Michtalik, Shaker M. Eid, Laura Wortman, MaryJane E. Vaeth, Charles F S Locke, Elizabeth Hoemeke, Raena Hariharan, Charles D. Callahan, James R. Ficke, Isabel Pimenta, Paul G. Auwaerter, Melinda E. Kantsiper, CONQUER COVID Consortium, Zishan K. Siddiqui

## Abstract

**Background:** Monoclonal antibody (mAb) treatment for COVID-19 has been underutilized due to logistical challenges, lack of access and variable treatment awareness among patients and providers. The use of telehealth during the pandemic provides an opportunity to increase access to COVID care.

**Methods:** This is a single-center descriptive study of telehealth-based patient self-referral for mAb therapy between March 1, 2021 to October 31, 2021 at Baltimore Convention Center Field Hospital (BCCFH).

**Results:** Among the 1001 self-referral patients, the mean age was 47, and most were female (57%) white (66%), and had a primary care provider (62%). During the study period, self-referrals increased from 14 per month in March to 427 in October resulting in a 30-fold increase. About 57% of self-referred patients received a telehealth visit, and of those 82% of patients received mAb infusion therapy, either onsite or at other infusion sites. The median time from self-referral to onsite infusion was 2 days (1-3 IQR).

**Discussion:** Our study shows the integration of telehealth with a self-referral process improved access to mAb infusion. A high proportion of self-referrals were appropriate and led to timely treatment. Incorporation of self-referral and telehealth for monoclonal antibody therapy led to successful timely infusions. This approach helped those without traditional avenues for care and avoided potential delay for patients seeking referral from their medical providers.

## Background

Since October 2020, six monoclonal antibody (mAb) treatments have received Emergency Use Authorization (EUA) from the Food and Drug Administration (FDA) for treatment of mild to moderate COVID-19 disease. At the time of their release, many predicted that the mAb infusion treatments would be exhausted in 1-2 weeks if used for all eligible patients. Some reports estimated about 30% of all COVID-positive patients would qualify for mAb therapy.[1][2] It has been suggested that mAb therapy has been underutilized in US.[3]

This underutilization has been attributed to logistical challenges related to timely testing, result notification, clinical evaluation, and referral for treatment within 7-10 days of symptom onset. Limited access to primary care providers (PCPs) and variable awareness among providers and patients about mAb treatment efficacy and eligibility also result in underutilization.[4][5] The dominance of the omicron variant has rendered the two most heavily used treatments useless. The explosive increase beginning in December 2021 of COVID-19 cases and the limited availability of the only effective mAb sotrovimab resulted in many centers handling monoclonal antibody therapy as a scarce resource.

An increase in the acceptability and use of telehealth during the pandemic has afforded the opportunity to assess COVID patients for appropriateness of mAb treatment remotely.[6][7] The Baltimore Convention Center Field Hospital (BCCFH) infusion center initiated a patient self-referral pathway with a telehealth visit to improve access and to address logistical challenges of timely referral. These visits then assessed eligibility for treatment, discussed risks and benefits with the patient, and scheduled the same or next day infusion, including transportation if needed. We describe the feasibility and implementation of integrated self-referral and telehealth service at the BCCFH mAb infusion center. We discuss the patient population who utilized this service.

## Methods

### Study design

This is a descriptive single-site study of the development, implementation, and operational experience of a patient self-referred telehealth service for monoclonal infusion treatment for COVID-19. The study was approved by the Johns Hopkins institutional review board (IRB).

### Location

This service was developed and housed at the Baltimore Convention Center Field Hospital (BCCFH) Monoclonal Infusion Site. BCCFH is a Maryland Department of Health disaster hospital two academic medical centers operate in Baltimore City.[8–10] In addition to inpatient care, BCCFH is Maryland’s largest mass COVID testing, monoclonal infusion, and mass vaccination site. This location does not provide in-person clinic-based ambulatory care (e.g., ambulatory primary care provider or specialist services).

### Time-period

We describe the development, implementation process and subsequent experience from the start of this service in March 1, 2021 until October 31, 2021.

### Data

Self-referral data was obtained from a database linked to a HIPAA (Health Insurance Portability and Accountability) compliant electronic web-based self-referral form. This patient-facing form was developed using Smartsheet software. This dataset provided data related to eligibility (clinical risk factors like obesity etc.), demographics, insurance type, referral source, date of symptoms onset, date of test positivity, date of tele-visit, type of telehealth visit, the outcome of telehealth visit, and date of infusion treatment. It was further updated by administrative and clinical staff with the status of referral and treatment. The development of the form is further described in the implementation section below.

### Analysis

The number of referrals, telehealth visits, infusion treatment and patient characteristics were summarized. Symptom-onset to self-referral, telehealth, and infusion treatment times were calculated. Additionally, self-referral to telehealth visit time, self-referral to treatment time, and telehealth visit to treatment time each were calculated. Patient characteristics of those who self-referred and had a telehealth visit were compared with those who self-referred and did not have a telehealth visit. Similarly, patients with audio-only visits were compared with video & audio telehealth visit. Amongst patients who had a telehealth visit, patient characteristics for those who received infusion were compared with those who did not.

JMP® Pro 16.0.0 was used for statistical analysis. The study was approved by the local institutional review board (IRB).

## Results

### Development and Implementation

1. Developing a self-referral stream:
  a. Increasing patient awareness about the treatment availability: A multimodal approach was employed to publicize the service among potential patients. Information about the mAb treatment and the process of self-referral was provided at the SARS-CoV-2 testing site at BCCFH. The BCCFH also partnered with the existing public health program of contact tracing through the state and local health departments. The contact tracing teams across the state were already tasked to reach individuals who tested positive for COVID. The BCCFH presented webinars to these teams to educate them about the mAb treatment, the barriers to access for patients, and BCCFH’s self-referral service for treatment. The contact tracers would direct eligible patients to BCCFH web-based self-referral forms and self-referral telephone line. They were given simple instructions to screen outpatients who were either less than 12 years of age or who had been ill for longer than 10 days. A script was provided for patient interaction to include information about treatment and the process of self-referral.
  b. Web-based and telephone self-referrals system: Patients had two options to complete the self-referral form: either by completing a web-based Smartsheet intake form or by calling the BCCFH telephone line so that BCCFH staff could complete the Smartsheet form on their behalf. The form itself was developed by BCCFH clinical leaders in collaboration with an institutional expert user and required minimal input from the institutional IT team in its development. Once content was finalized, the form was built within a day. The form captured the patient’s demographic information, health history, and COVID clinical history. A completed form transmitted information directly into a spreadsheet database. The spreadsheet coding allowed the team to identify and prioritize patients based on criteria such as time since symptom onset, and clinical risk factors.
2. Intake, screening and scheduling a telehealth visit : All patients were contacted by the intake team to review patient information and schedule a telehealth appointment. The intake occurred during the same phone call for patients who called to self-refer. The intake team included medical assistants, social workers, and nurses supervised by the medical director. Due to staffing constraints at the BCCFH, most intakes were performed by the social workers. Equipped with a script and a list of frequently asked questions and answers, the intake team could provide the basic information about the medication and infusion process. The intake team could screen for in eligible patients under the supervision of the medical director who was available either in the same working space or via a HIPAA compliant instant communication application. Patients evaluated by medical assistants or social workers deemed ineligible if past the 10-day eligibility period used during the study period, but otherwise, were scheduled for a telehealth visit. Patients assessed by nurses, who had been trained on the specifics of the eligibility for mAb treatment, were given preliminary determinations about their eligibility for mAb-based on underlying health conditions. Although some patients were further screened out this way, a telehealth visit was scheduled for ineligible patients if they still requested it. All patients who met EUA criteria for mAb treatment were offered a telehealth visit with a provider. The intake team scheduled the telehealth visit in the electronic medical record (Epic). The intake team also provided patients instructions to access Zoom and the link to the telehealth appointment via email. Patients with barriers to accessing zoom were scheduled for a telephone-only appointment. During periods of lower volume, one intake staff person managed referrals and built schedules of telehealth visits for up to two providers. During peak times, the team consisted of two staff people processing referrals and building schedules for three providers.
3. Telehealth visits with provider BCCFH was able to build this service without any prior ambulatory services. Telehealth model allowed rapid creation of this service line without making investments in a physical location, supplies, support staff. This service was developed by redeploying an existing pool of providers at BCCFH from its inpatient, vaccine, and testing service. They received additional training about mAb eligibility criteria, telehealth visits, care coordination, and scheduling. Eligible patients able and willing to get treatment at BCCFH were scheduled for an infusion treatment during the visit. This led to immediate patient scheduling, rather than waiting for another scheduling call. When patients preferred, or when earlier appointments were available elsewhere, referrals were made to outside infusion sites, obviating the need for patients to be seen by another provider for a referral to these sites. The standard time allotted to a provider’s telehealth visit was 30 minutes. The providers also instructed the patients on preparing for the visit (e.g., hydration) and provided directions to the site. Patients received an email about appointment, location, and instructions to prepare for the day of infusion. There was no direct cost to the patient irrespective of their insurance status, although insurance was billed if available.

### Operational Experience

The self-referral and telemedicine services were established in March 2021, but self-referrals volumes remained low until July 2021, after which the self-referral volume increased by more than 50% each subsequent month. There was a similar increase in tele-visits and infusions (Figure 1). This rise coincided with increased rates of infection and hospitalization in the state.[11] During the study period, 1001 self-referrals were received. Self-referred patients had a mean age of 47, were more frequently female (57%), White (66%), English speakers (98%), non-Baltimore City residents (85%) and reported having a PCP or continuity care provider (62%). Most patients learned about the self-referral service from the state health department contact tracers (60%). Others learned about the service from family or friends (12%) and 11% learned from ED/Urgent care providers or other health care personnel (Table 1).

**Figure 1:**
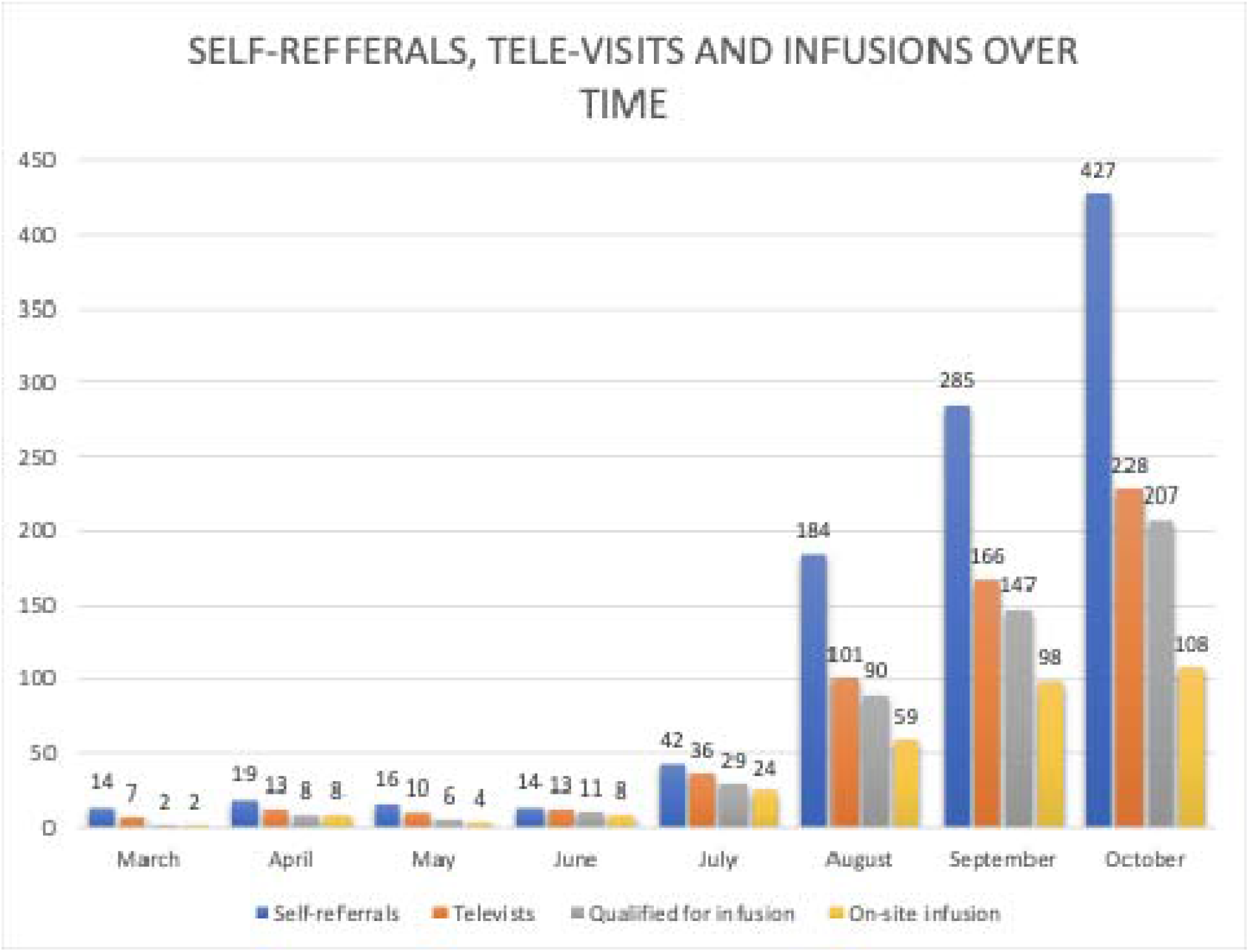
Self-referrals, subsequent tele-visits and infusions from May 2021 to October 2021 About 30-fold increase in self-referrals and a 50-fold increase in on-site mAb infusions between march 2021 to October 2021

**Table 1:** Self-referral Patient Characteristics and Demographics.

| **Characteristics** | **n=1001** |
| --- | --- |
| **Age (Mean)** | **47 years** |
| **Female** | **57%** |
| **Race** |  |
| **White** | **66%** |
| **African American** | **22%** |
| **Latin** | **6%** |
| **Other** | **7%** |
| **Tele-visit type** |  |
| **Audio only** | **22%** |
| **Audio and Video** | **78%** |
| **Preferred Language** |  |
| **English** | **98%** |
| **Spanish** | **2%** |
| **Other** | **0.1%** |
| **Reported Having a PCP/Continuity Provider** | **62%** |
| **Baltimore City Residence** | **15%** |
| **Learned about self-referral service from:** |  |
| **Maryland Department of Health\*** | **60%** |
| **Baltimore City Health Department** | **4%** |
| **Family/Friend** | **12%** |
| **Urgent Care/ED** | **3%** |
| **Other Healthcare Source** | **8%** |
| **Web Search** | **6%** |
| **Other** | **7%** |
| **\*Contact tracing program of Maryland Department of Health** |  |

Of the 1001 self-referred patients, 50% received mAb treatment at BCCFH or other sites. During the intake process, 14% of the self-referred patients were unreachable, 10% sought treatment elsewhere, 7% declined treatment, and 5% were ineligible for treatment. These patients were not scheduled a tele-visit appointment. Three percent were scheduled for infusion without a telehealth visit because a provider referral was also received for these patients. Overall, 427 (43%) of self-referrals did not have a tele-visit. Among these, 574 (57%) individuals were scheduled for a tele-visit. Of these, 290 (50%) received infusion on site, and an additional 32% were referred off-site for treatment, either because of their proximity to those centers or to enable infusion sooner than possible at our site. The remaining 18% of the patients who had a tele-visit did not receive an infusion, mainly because they were ineligible (8%) or declined treatment (6%) (Figure 2).

**Figure 2:**
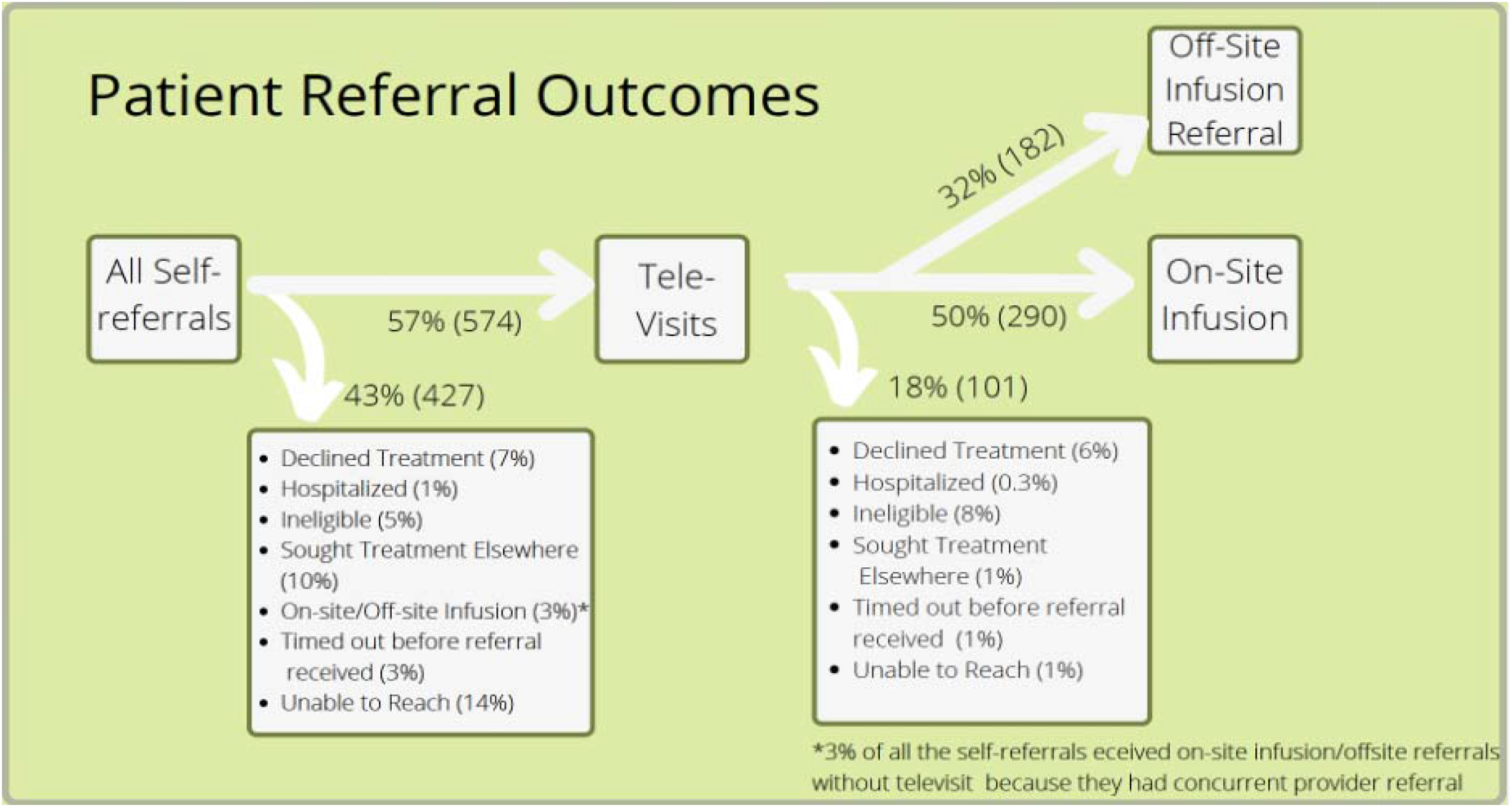
Distribution of patients by outcomes after self-referral

**Figure 3:**
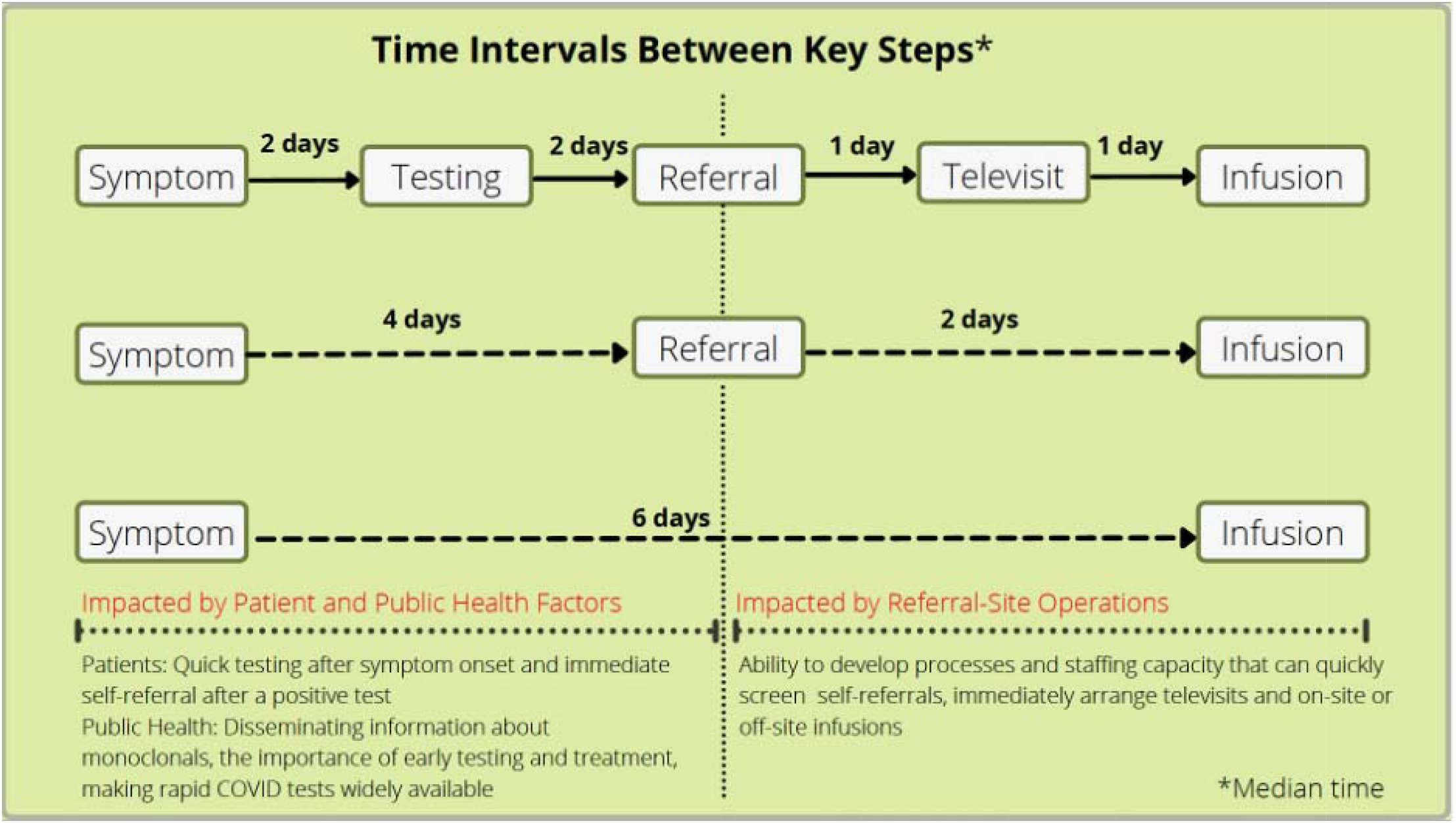
Median time intervals between key steps from symptom onset to monoclonal infusion therapy among self-referred patients

The median time from symptom onset to onsite infusion was 6 days (5-8). Self-referred patients obtained a COVID test after a median of 2 days (1-3) and self-referred after a median of 4 days (3-6) from onset of symptoms. Patients had a median 1 day (0-1) wait from self-referral to tele-visit and 2 days (1-3) between self-referral and onsite infusion. An estimated 57% of the patients received telehealth visits the same day as referral, 83% within 1 day of self-referral, and 72% of the patients received onsite infusion within 2 days of referral. 6% of the patients received onsite infusion the same day as self-referral and 13% received infusion the same day as tele-visit.

Type of tele-health visit (video& audio vs. audio only) was not associated with increased likelihood of qualifying for infusion (OR 0.7, 95% CI 0.4-1.2), or onsite infusion (OR 1.1, 95% CI 0.8-1.7). Among self-referred patients, those who received tele-health visits were no different from those who did not receive tele-health visits on race, gender, Baltimore city residency, where they learned about the self-referral service and reported having a PCP, symptom to test time and symptom referral time (Table 2). Among the patient who had a tele-visit, those who did not receive infusion were compared to those who received infusion (onsite or off-site). There were no differences regarding race, gender, Baltimore city residency, where they learned about the self-referral service, reported having a PCP, symptom to test time, symptom onset to referral time, self-referral to tele-visit time.

**Table 2.** Supplemental table

| Characteristics | Had Tele-visit (574) | Did not have Tele-visit (427) | P-value | Had infusion (on-site or off-site) (472) | Did not have infusion (101) | P-value |
| --- | --- | --- | --- | --- | --- | --- |
| Age (Mean) | 47 | 47 | 0.7 | 49 | 40 | <0.0001 |
| Female | 57% | 58% | 0.7 | 58% | 61% | 0.6 |
| Race |  |  |  |  |  |  |
| White | 62% | 71% | 0.06 | 64% | 58% | 0.4 |
| African American | 24% | 19% |  | 22% | 26% |  |
| Latin | 6% | 5% |  | 7% | 5% |  |
| Other | 8% | 6% |  | 7% | 10% |  |
| Reported Having a PCP/Continuity Provider | 61% | 63% | 0.5 | 63% | 57% | 0.3 |
| Baltimore City Residence | 15% | 16% | 0.7 | 16% | 15% | 0.9 |
| Learned about self-referral service from: |  |  |  |  |  |  |
| Maryland Department of Health | 59% | 61% | 0.1 | 55% | 66% | 0.2 |
| Baltimore City Health Department | 4% | 4% |  | 5% | 4% |  |
| Family/Friend | 13% | 12% |  | 14% | 11% |  |
| Urgent Care/ED | 2% | 3% |  | 3% | 1% |  |
| Other Healthcare Source | 7% | 9% |  | 8% | 4% |  |
| Web Search | 5% | 6% |  | 4% | 8% |  |
| Other | 10% | 5% |  | 11% | 7% |  |
| Symptom to Test Time (days) | 2.1 | 2.2 | 0.9 | 2.2 | 1.9 | 0.2 |
| Symptom to Self-referral Time (days) | 4.6 | 4.8 | 0.3 | 4.9 | 4.5 | 0.2 |
| Symptom to Tele-visit Time |  |  |  | 5.4 | 5.6 | 0.6 |
| Self-referral to Tele-visit Time |  |  |  | 0.8 | 0.8 | 1.0 |

Those who received infusion onsite, compared to those who were sent for infusion off-site were less often referred by the Maryland Department of Health and were more often Baltimore-City residents and referred by Urgent Care/ED and the Baltimore City health department.

## Discussion

In this single-center descriptive study, we demonstrate the feasibility and experience of establishing a telehealth-based patient self-referral service for COVID-19 monoclonal antibody infusion treatment. This mechanism addresses the unmet needs as evidenced by high utilization of the service. The telehealth model allowed rapid creation of the service without the typically significant investments needed for onsite care. About 50% of the self-referred patients received successful infusion treatment demonstrating that a substantial proportion of patients appropriately self-referred. During the study period, self-referrals increased from 14 per month in March to 427 in October resulting in a 30-fold increase. The number of infusions for self-referral pathway increased from 2 to 104 per month during the same period, marking a 50-fold increase.. A consistent increase in the volumes of self-referral and telehealth visits over time were likely related to increased awareness of the service and increased disease prevalence in the community. BCCFH was especially successful in partnering with the existing public health programs, as 60% of the self-referral patients were informed about this service through the state’s contact tracing program.

This appears to have mitigated some barriers and expanded access for ambulatory patients seeking care for non-severe COVID-19. Almost two-thirds of the self-referred patients report having continuity providers, suggesting that timely access for COVID care remains an issue in this population when an urgent visit is needed to meet a narrow window of eligibility for mAb therapeutics. Additionally, about 11 % of patients were directed to this service through ED, urgent care and other healthcare services, perhaps because they were overstretched, or the test results came back after this visit. The telemedicine-based self-referral process also provides an alternative pathway for timely access to patients who do not have access to primary care physicians and may potentially assist reducing crowding in emergency departments and urgent care locations during surges. [13–17]. Additionally, telemedicine visits for COVID-19 patients allow for better patient comfort, decreased chances of disease transmission, reduced healthcare expenditure for space, supplies and staff compared to in-person care clinic.[18]

A mAb focused telehealth service may also support existing primary care providers as same-day appointments with primary care providers may be challenging.[12]

Contrary to hesitations raised toward self-referral service in the past, we found that a high percentage were eligible and received mAb treatment (56%).[16] Additionally, more than 80% of patients who had a telehealth visit were scheduled for treatment. A patient-centered approach was utilized during scheduling as 32% of the patients who had telehealth visits were scheduled at outside sites for convenience or timely care.

Monoclonal antibody therapy is a time-sensitive treatment since patients were eligible for 10 days from the onset of symptoms during the study period, and have the great impact if administered early in the infection.[19] The median time from symptom onset to infusion was 6 days, similar to other centers with mainly provider-referred infusions.[12,14,19] Time from referral to infusion treatment is affected by the infusion site operations and is a key performance metric for an infusion center. Our study shows telehealth model provides an efficient method for evaluating these patients for eligibility. The 2-day median time from self-referral to infusion at our center is similar to other centers, including provider referrals, patient self-referrals, new diagnoses, and treatment in the emergency department and inpatient units.[13–17]

Our case study has a few limitations. Our center has a unique organizational structure as a public-private partnership. This allowed us to collaborate with the State’s contact tracing system to develop a referral system and allowed us to use the capabilities of academic centers to innovate. Hence, our process may not be generalizable. However, the model may still be used to connect with patients at other COVID self-test sites. Second, we could not track and report specifically about the patients who used telephone service to self-refer. The intake team estimated that it represented less than 5% of self-referrals hence our measures of process and outcomes may not apply to them. They may be better described by other centers that see a large percentage of these patients.

## Conclusion

A sustained public health emergency such as the COVID pandemic has resulted in new models of care. Unlike customary healthcare ambulatory practice, patients were able to gauge the need for COVID testing, self-refer to testing sites based on their knowledge of self-directed diagnostic testing. They directly receive, interpret, and act on the results. Self-referral infusion service is a logical extension of the patient self-driven public health response to the pandemic. This is especially useful when the primary care providers or specialists cannot see patients in the very short time frame needed to meet the eligibility window for the treatment. Our case study shows that self-referral for mAb treatment integrated with telehealth is feasible and results in timely access to treatment. Partnering with multiple agencies responsible for testing, contact tracing, and patient awareness about the service would be key for sustained utilization. Operational development should focus on creating referral streams, screening protocols and staff capability. Key quality metrics like referral to infusion time should be monitored and operations adjusted to keep this indicator low. Self-referred telehealth service will remain relevant in shifts in ambulatory treatment from infusions to oral therapeutics.

## Data Availability

Data is not publically available.

